# Assessing the relationship between measures of healthcare access and functional limitations among individuals with Chronic Obstructive Pulmonary Disease (COPD)

**DOI:** 10.1101/2021.12.30.21268572

**Authors:** O J Adeyemi, A A Arif, R Paul

## Abstract

**Objectives:** Functional limitation from COPD manifests more from physical rather than respiratory impairment. To what extent health access affects the functional limitation among individuals with COPD is yet to be known. This study aims to assess the relationship between healthcare access and functional limitations among individuals with COPD.

**Study Design:** Retrospective analysis of a cross-sectional population-based survey

**Methods:** This study pooled 11-year (2008 - 2018) data from the Integrated Public Use Microdata Series - National Health Interview Survey (IPUMS-NHIS). We restricted the data to respondents with self-reported COPD, aged 40 years and older. The independent variables were sociodemographic and behavioral characteristics. The exploratory variables were measures of healthcare access – healthcare coverage, delayed appointment, affordable care, and a usual place for care. The outcome variable was the presence or absence of functional limitations.

**Results:** The age, race, educational attainment, marital status, smoking status, and poverty-income ratio had a significant association with functional limitation (p<0.001) We found statistically significant associations between functional limitation and healthcare coverage, delayed appointment, affordable care, and a usual place for care. Poverty modified the relationship between functional limitations and the four measures of healthcare access, with the odds of functional limitation increased among the poor with no healthcare coverage, delayed appointment, unaffordable care, and no usual place for care.

**Conclusions:** A strong relationship exists between the quartet of healthcare coverage, delayed appointment, affordable care, and usual place for care and self-reported functional limitation among individuals with COPD. Poverty was an effect modifier, with the odds of functional limitation worse among the poor.

## Introduction

In the United States (U.S.), COPD is the fourth leading cause of death ^1^. Hospitalizations following acute exacerbations of COPD account for $18 billion in direct hospital costs ^2^. Functional limitation from COPD, a measure of health decline and a predictor of worse health outcomes ^3,4^, manifests more from physical rather than respiratory impairment. Individuals with COPD, aside from having decreased lung function and occasional exacerbations, develop progressive decrements in strength and mobility ^5^. Worse still, COPD is more often complicated by comorbidities, which further exacerbates the functional limitation, worsens the patients’ health status, and increases the need for health care utilization, hospitalization, and expenses ^6^.

Despite the burden of COPD, it is a disease that is rarely viewed through the lens of health access. Consistent across most COPD-related studies, the odds of COPD occurrence is worse among individuals with low educational attainment, the poor ^7-10^, and the Blacks and Hispanic population ^5,7,11^. Additionally, Blacks with COPD have lower healthcare utilization compared to Non-Hispanic Whites ^12^. Chronically ill patients without insurance, including those with COPD, are more likely not to have regular visits to a healthcare professional or have a proper site for care aside from the emergency department ^13,14^. Further, poverty is associated with increased odds of functional limitation from any disease ^3,15^.

To what extent health access affects the functional limitation among individuals with COPD is yet to be known. For a disease that affects a substantial proportion of the U.S. population, the impact of healthcare access among people of color, race, residence, socioeconomic, and health insurance status is not only desirable but necessary. Therefore, this study sets out to assess the relationship between healthcare access and functional limitations among individuals with COPD. We hypothesize that the absence of health coverage, lack of a usual place for care, affordable care, and the presence of delayed appointments would be associated with increased odds of functional limitation among individuals with COPD. Secondly, we hypothesize that poverty is an effect modifier, that increases the odds of functional limitation from healthcare access measures among the poor.

## Materials and Methods

In this study, 11-year data (2008 - 2018) from the Integrated Public Use Microdata Series - National Health Interview Survey (IPUMS-NHIS) were pooled and analyzed ^16^. The National Health Interview Survey (NHIS) represents a cross-sectional survey of a sample population of the civilian, non-institutionalized United States population ^17^. It is the longest national survey in the U.S., sampling approximately 35,000 households and 87,500 individuals yearly ^17^. The NHIS uses a multi-stage cluster sampling method to select households and individuals across all U.S. states and the District of Columbia ^17^. The average yearly response rate is about 70% ^18^. The interview questionnaires majorly obtain information across several domains, which include household, family, sample adult, sample child. Additional details about the survey and data collection methods are available online at https://www.cdc.gov/nchs/nhis/about_nhis.htm. There had been changes in the coding and wording of some of the NHIS survey items across time. The Integrated Public Use Microdata Series (IPUMS) harmonizes these changes in a user-friendly way that facilitates temporal pooling of the NHIS data, observation of the consistency and comparability of survey item across time, and the response, and the yearly response counts of each survey item ^19^. Several studies have used the IPUMS-NHIS as a source of NHIS data ^20-22^.

Across the eleven years (N= 1,031,158), we selected individuals with COPD (n=18,508). We defined cases of COPD by responses to the questions: “Have you ever been told by a doctor or other health professional that you had …Emphysema?” and “During the past 12 months, have you been told by a doctor or other health professional that you had …Chronic bronchitis?” Previous studies have used these two questions to define COPD, including a third question on ever being told that they have COPD ^23,24^. The IPUMS-NHIS does not have this third defining variable. Further, we restricted the data to respondents ages 40 years and older (n= 15,558), consistent with previous studies ^25,26^. We used this cut-off since COPD prevalence increases from ages 40 years and older ^26,27^. Independent variables extracted from the dataset included socioeconomic variables such as age, gender, race/ethnicity, educational level, marital status, smoking status, and poverty level. These variables were used as a priori confounders. We performed a listwise deletion for missing variables in the outcome variable since the missing counts were less than 1% ^28^. Hence, the final dataset comprised of 15,216 respondents, with 8,522 having functional limitation and 6,694 without functional limitation (Figure 1).

**Figure 1:**
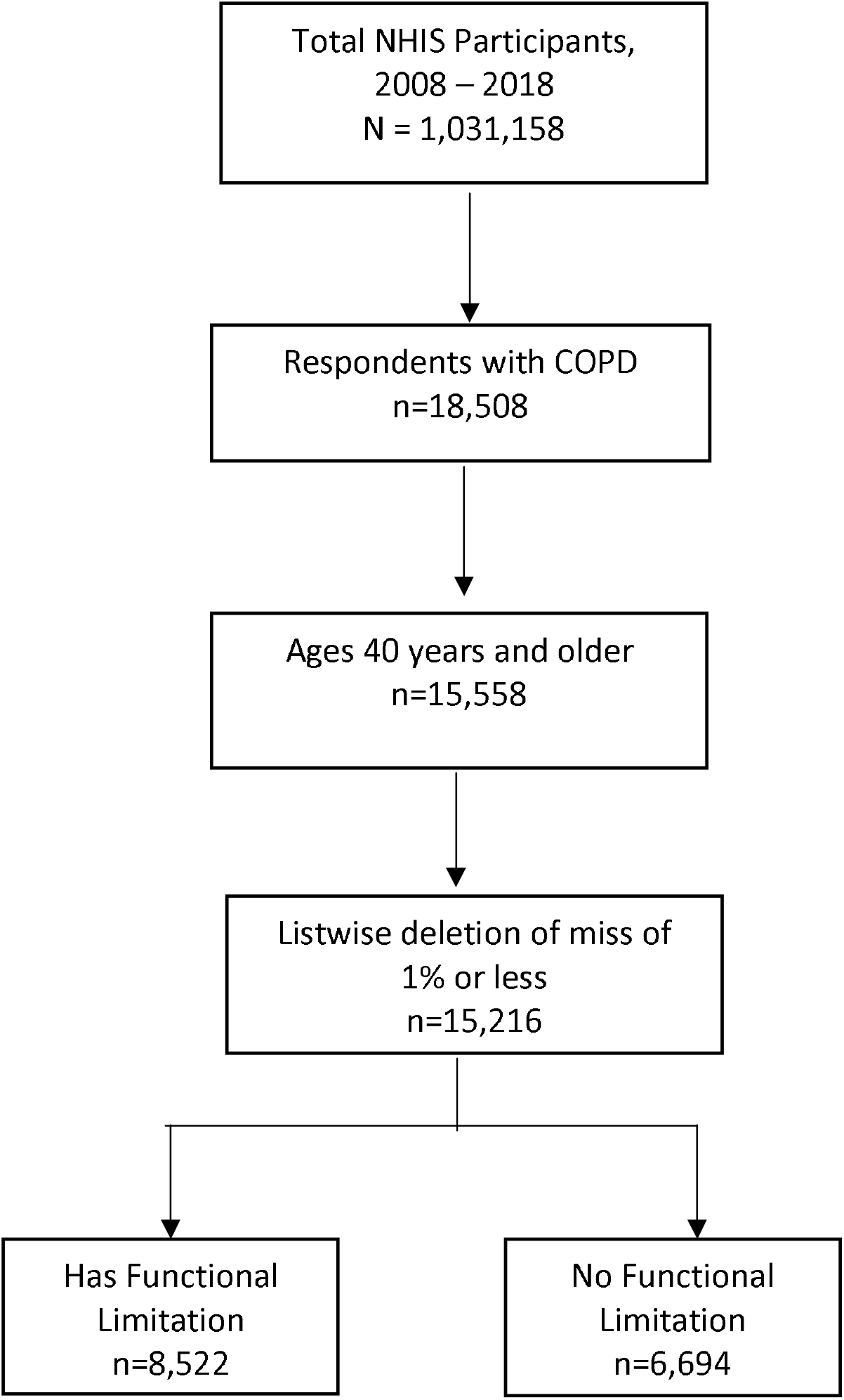
Data selection steps

The outcome variable of interest was functional limitations, a computed variable that measured whether the respondent had any activity limitation. Specifically, the variable was recoded (by IPUMS-NHIS) from responses to the questions that assessed if the respondent is limited from any physical, mental, or emotional problems, that makes the respondent need help with routine needs, prevent the respondent from working at a job or business, have difficulty walking without special devices, or limited in any other way. The responses were coded as a four-point categorical variable: limited in any way, not limited/unknown, not limited in any way, or unknown if limited. We defined functional limitations as either limited in any way or not limited. Respondents who do not know if they had limitations (n=10) were excluded from the study.

For this study, the main explanatory variables were four measures of healthcare access: health coverage, delayed appointment, affordable care, and a usual place for care. We defined health coverage by the response to the question if the respondent lacks health insurance coverage at the time of the interview. Also, we defined delayed appointments by the question, “have you delayed getting care in the past 12 months because you could not get an appointment soon enough?” We defined affordable care by the question asking if, in the last 12 months, the respondents needed healthcare but could not afford it. We defined a usual place for care by the response to the question if the respondents had a usual place for medical care. For this study, the poverty level was used as an interaction variable since the presence plays a role in health-seeking behavior and access to health ^29^. Respondents whose family income was less than the 250% threshold of poverty were classified as poor. The respondents, at or above the poverty threshold, were classified as not poor.

Data were analyzed using STATA version 14. Descriptive parameters were reported in tables, and the bivariate relationship between the functional limitation and the sociodemographic and explanatory factors were assessed using chi-square statistics. We visualized the trend of functional limitation across time to evaluate if there were significant changes across the years. In answering the first hypothesis, we performed a univariate and multivariate logistic regression analysis to estimate the odds of functional limitation and healthcare access measures. For each of the four main explanatory variables, we controlled for potential confounders – age, gender, race/ethnicity, educational level, marital status, smoking status, and poverty level. We created a fifth model that assessed all four explanatory variables while controlling for potential confounders.

Further, to answer the second hypothesis, we assessed the relationship of functional limitation and the explanatory variables using the poverty level as an interaction variable while controlling for the other sociodemographic and behavioral factors. We reported the adjusted odds and 95% confidence interval of each explanatory variable, stratified by poverty level. We applied survey weights to adjust for the complex sampling design of the NHIS. Hence, the analytical procedure involved the use of survey-weighted frequencies and survey-weighted logistic regression. Since we pooled eleven years of data, we calculated the weights by dividing the person weight variable – an IPUMS-NHIS computed variable, by 11 ^30,31^.

## Results

Between 2008 and 2018, a total of 15,216 respondents with COPD, representing 3,983,061 non-institutionalized Americans, participated in the survey. About 55% of the respondents, representing 2,198,650 Americans with COPD, reported that they had functional limitations. More than 59% of the respondents were 60 years or older. The majority were female (64%), non-Hispanic White (80%), and had high school or less education (53%). About 70% of the respondents were current or past smokers, and 20% were living below the poverty level. The proportion of respondents with functional limitation increased with increasing age (p<0.001). A bivariate analysis (chi-square) showed that race (p<0.001), educational attainment (p<0.001), and marital status (p<0.001) had significant associations with functional limitations. The smoking status and poverty income ratio index had a statistically significant relationship with functional limitations, with 76% of current or past smokers having functional limitations (p<0.001) and 63% of respondents at or above the poverty income index having functional limitations (p<0.001). Furthermore, health insurance coverage, delayed appointment, affordable care, and the usual place for care were statistically significant with functional limitations (p<0.001) (Table 1).

**Table 1:**
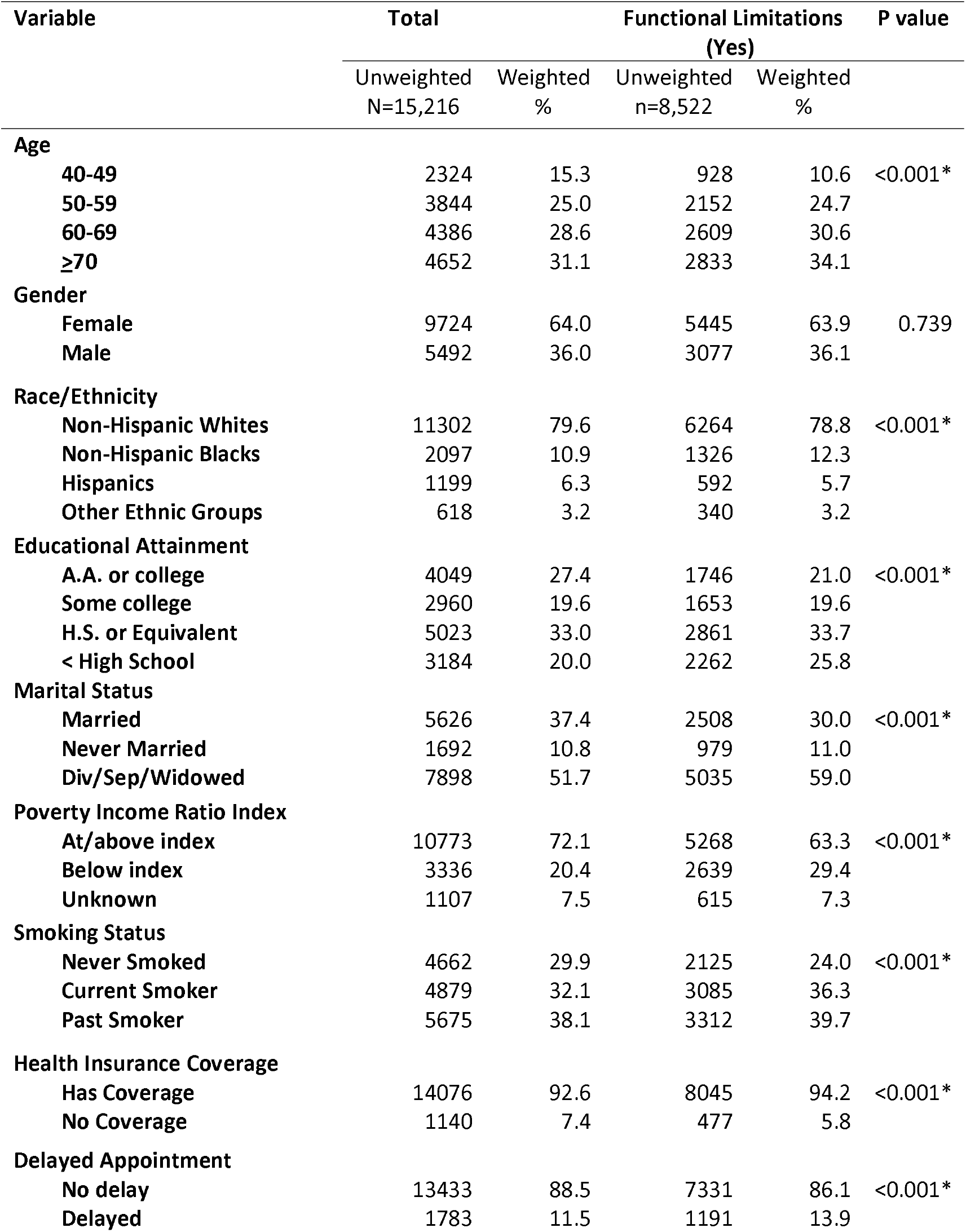
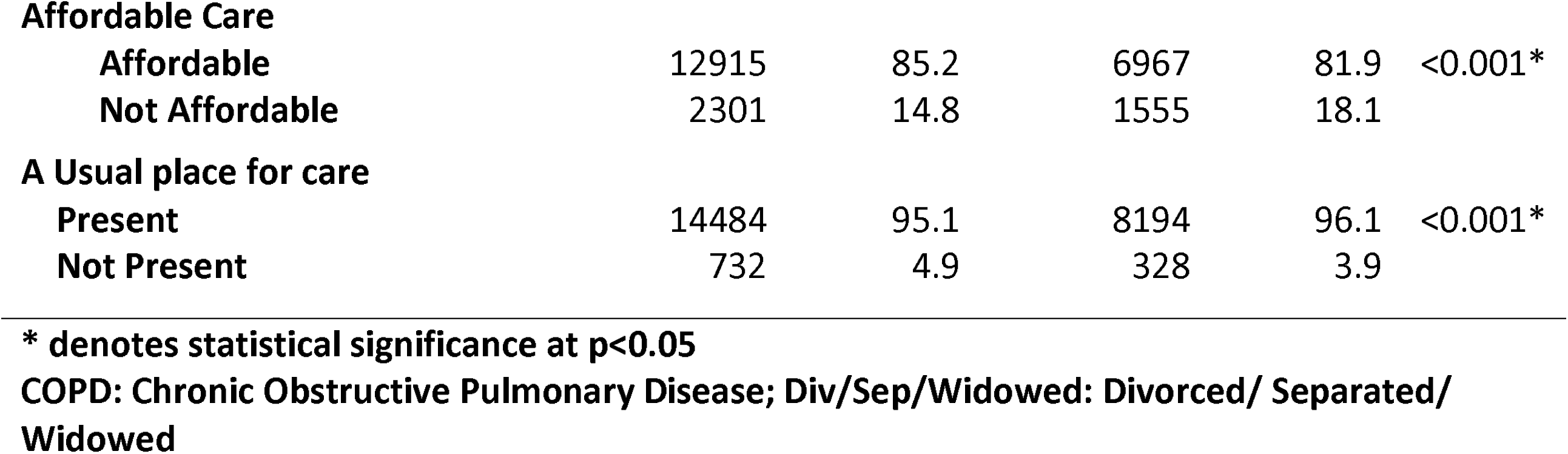
Frequency distribution of sociodemographic factors and explanatory variables of NHIS respondents with COPD stratified by functional limitation (N=15,548), NHIS 2008-2018

Consistently across time, the weighted proportion of individuals with COPD ranged between 51 - 57%, and there was no significant decline across time (Figure 2A). Additionally, non-Hispanic Blacks had a higher weighted proportion of functional limitation compared to non-Hispanic Whites. Among the Hispanic population, there was a gradual increase in the weighted proportion of individuals with functional limitations from COPD (Figure 2B).

**Figure 2:**
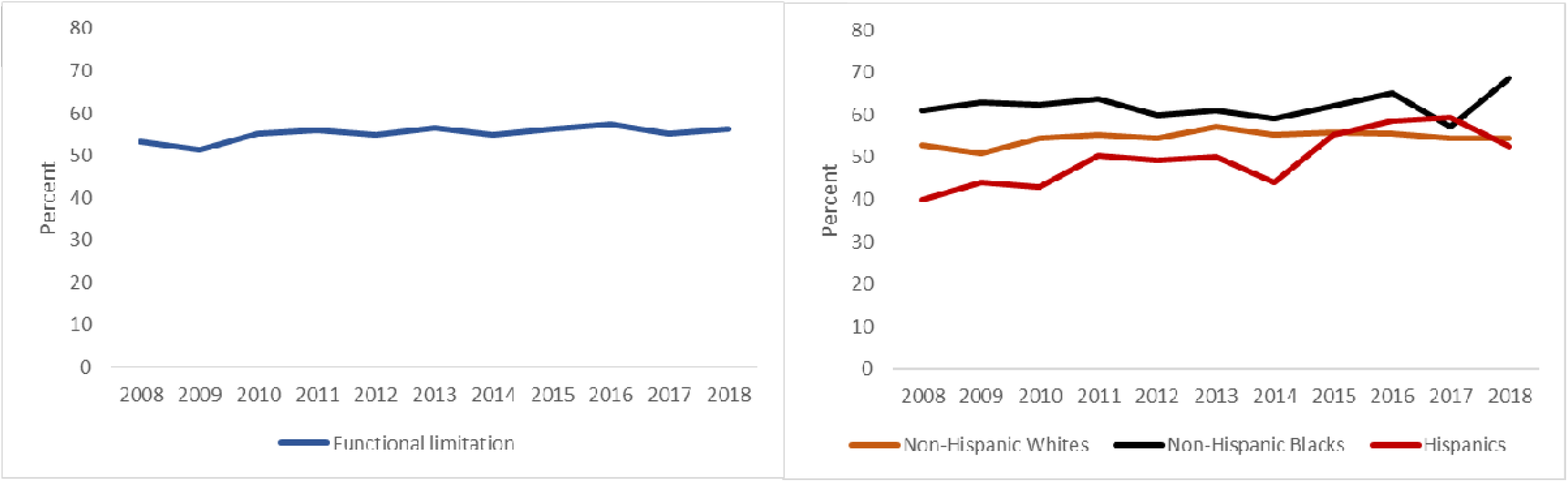
Proportion of functional limitation among respondents with Chronic Obstructive Pulmonary Disease (A) across the eleven-year period (2008 - 2018) and (B) by race/ethnicity.

We performed a univariate logistic regression analysis to assess the relationship between the explanatory variables and functional limitations from COPD (Table 2). In the unadjusted model, respondents without health insurance were 42% less likely to have functional limitation (Odds Ratio (OR): 0.58; 95% CI: 0.50 – 0.68), while those without a usual place for care were 38% less likely to have functional limitations (OR: 0.62; 95% CI: 0.52 – 0.74). However, respondents with delayed appointments were 1.7 times (95% CI: 1.52 – 1.92) more likely to have functional limitations, and those who cannot afford care were 1.8 times (95% CI: 1.64 – 2.06) more likely to have functional limitations.

**Table 2:** Univariate and multivariate logistic regression analysis of the association of functional limitation and the explanatory variables adjusted for sociodemographic and behavioral characteristics using NHIS data: 2008 - 2018.

| Variable | Univariate Analysis | Model 1 | Model 2 | Model 3 | Model 4 | Model 5 |
| --- | --- | --- | --- | --- | --- | --- |
|  | Unadjusted OR (95% CI) | Adjusted OR (95% CI) | Adjusted OR (95% CI) | Adjusted OR (95% CI) | Adjusted OR (95% CI) | Adjusted OR (95% CI) |
| Health Insurance Coverage |  |  |  |  |  |  |
| Has Coverage | Ref | Ref |  |  |  | Ref |
| No Coverage | 0.58(0.50-0.68) | 0.55(0.46-0.66) |  |  |  | 0.43 (0.35-0.53) |
| Delayed Appointment |  |  |  |  |  |  |
| Not Delayed | Ref |  | Ref |  |  | Ref |
| Delayed | 1.71 (1.52-1.92) |  | 2.06 (1.81-2.35) |  |  | 1.88 (1.65-2.14) |
| Affordable Care |  |  |  |  |  |  |
| Affordable | Ref |  |  | Ref |  | Ref |
| Not Affordable | 1.84 (1.64-2.06) |  |  | 1.98 (1.72-2.27) |  | 2.51 (2.14-2.93) |
| Usual place for care |  |  |  |  |  |  |
| **Present** | Ref |  |  |  | Ref | Ref |
| **Not present** | 0.62 (0.52-0.74) |  |  |  | 0.55 (0.46-0.67) | 0.57 (0.46-0.69) |

All models are adjusted for age, gender, race, educational attainment, smoking status, and poverty-income ratio.

Further, we conducted multivariate logistic regression analysis for each of the four explanatory variables (models 1 - 4) and a final model (model 5) containing all four explanatory variables, while controlling for age, gender, race, educational attainment, marital status, smoking, and the poverty-income ratio at each step (Table 2). Prior to this step, we established that there was no multicollinearity in the four explanatory variables and the poverty-income ratio variable - all five variables had weak correlations with each other (result not shown). We found that respondents without health insurance coverage (Adjusted OR (AOR): 0.55; 95% CI: 0.46 – 0.66) and a usual place for care (AOR:0.55; 95% CI: 0.46 – 0.67) were 45% less likely to have functional limitations. Functional limitation was associated with two-fold increased odds of having delayed appointment (AOR: 2.06; 95% CI: 1.81 – 2.35), and unaffordable care (AOR: 1.98; 95% CI: 1.72 – 2.27). The pattern of association did not change in the final model (model 5) with functional limitation significantly associated with reduced odds of not having health insurance (AOR: 0.43; 95% CI: 0.35 – 0.53) or a usual place for care (AOR: 0.57; 95% CI: 0.46 – 0.69) and significantly associated with increased odds of delayed appointment (AOR: 1.88; 95% CI: 1.65 – 2.14) and unaffordable care (AOR: 2.51; 95% CI: 2.14 – 2.93).

We found that the poverty-income ratio was an effect modifier in the relationship of functional limitation and health coverage, delayed appointment, affordable care, and a usual place for care (Table 3). Respondents who were poor and had no health coverage were 1.4 times more likely to have functional limitations (95% CI: 1.08 – 1.85). Paradoxically, respondents who were not poor but had no health coverage were 31% less likely to have functional limitations (AOR: 0.69; 95% CI: 0.56 – 0.85). When we assessed the interaction between delayed appointment and functional limitation, the odds ratio between the poor and the not-poor quadrupled. Respondents who were poor with delayed appointments were about eight times more likely to have functional limitations (AOR: 7.57; 95% CI: 5.37 – 10.66). However, respondents who were not poor but have delayed appointments were two times more likely not to have functional limitations (AOR: 2.07; 95% CI: 1.78 – 2.40). Similarly, respondents who could not afford care and were poor were 4.5 times more likely to have functional limitations (95% CI: 3.55 – 5.70), while respondents who could not afford care but were not poor were 2.3times more likely to have functional limitations. There exists a paradox in the effect of poverty on the relationship between usual place for care and COPD-related functional limitations. Respondents who were poor and had no usual place for care were 1.6 times more likely to have functional limitation (95% CI: 1.17 – 2.16) while respondents who were not poor were about 39% less likely to have functional limitation (AOR: 0.61; 95% CI: 0.48 – 0.77) (Table 3).

**Table 3:** Interaction Analysis Between the Odds Ratio of the Exposure variables and the Poverty-Income Ratio in estimating the odds of functional limitation among individuals with COPD using NHIS data: 2008-2018

| Variables | Poverty Income Ratio (OR (95% CI))** |  |
| --- | --- | --- |
|  | Not Poor | Poor |
| No Health Coverage | 0.69 (0.56 - 0.85) | 1.42 (1.08 - 1.85) |
| Delayed Appointment | 2.07 (1.78 - 2.40) | 7.57 (5.37 - 10.66) |
| Unaffordable Care | 2.27 (1.93 - 2.65) | 4.49 (3.55 - 5.70) |
| No Usual place for care | 0.61 (0.48 - 0.77) | 1.59 (1.17 – 2.16) |

All odds ratios adjusted for age, gender, race, educational attainment, marital status, and smoking status
\*\*Interaction odds ratios were calculated by first including an interaction term of exposure x poverty-income ratio (PIR) index and then exponentiating the sum of the coefficients of PIR(effect modifier), exposure, and the interaction term.

## Discussion

Consistent with our first hypothesis, delayed appointment and unaffordable care were associated with functional limitation. However, the lack of health coverage and the lack of a usual place for care were paradoxically associated with reduced odds of functional limitation. Our second hypothesis unraveled this paradox as the lack of coverage and a usual place for care was associated with functional limitation only among those who were poor and not among those who were not poor. Additionally, consistent with our second hypothesis, poverty modified the effect of delayed appointments and unaffordable care, and functional limitation. Compared to those who were not poor, the odds of functional limitation among the poor with a delayed appointment and unaffordable care increased by a four-fold and two-fold, respectively. The odds of functional limitations were higher among the individuals who were poor, had COPD, and were without health insurance. This association was exactly the opposite among those who were not poor yet have COPD and were without health insurance.

Health insurance coverage is a barrier to patient care ^32^, and the lack of health insurance among adults in the United States increases their odds of dying ^33^. Furthermore, health insurance availability and expansion to poorer counties have been associated with a positive self-rated health assessment ^34^. Other authors have alluded that health insurance status is one of the perceived barriers to patient’s care ^14^, and its impact is reflected in reduced access to needed health among children ^35^, adults and the elderly ^14^. Among individuals with COPD, the influence of lack of health insurance on health outcomes is evident among the poor. This study highlights the need for health insurance coverage, especially for the poor population ^32^.

In our study, individuals with COPD who were not able to afford healthcare services were more likely to have functional limitations, with poverty increasing the odds by two-folds. Earlier studies have reported that poverty worsens the odds of being able to afford healthcare services ^36^. With the cost of managing COPD in the United States one of the highest in the world ^37,38^, individuals with COPD are more likely to use the health facility to treat other illness rather than acute exacerbations of COPD ^14^.

Our study found that the odds of functional limitation were associated with lack of a usual place for care, with the poor having increased odds of functional limitation, while those who were not poor had reduced odds. A possible explanation is that individuals who are poor rationalize their health needs ^37,38^. Additionally, individuals with COPD have a lower household income compared to the general population ^5^ and they prioritize presenting at the emergency rooms in acute exacerbations than attending regular clinic visits ^14,39^. Andersson and colleagues ^14^ noted that individuals with COPD were admitted for COPD-related illnesses more towards their end of life, estimating that about 3% of patients with COPD were never admitted till their demise.

Our study found that individuals with COPD who had delayed appointments were more likely to have functional limitations, with poverty generating seven-fold increased odds of functional limitation. Indeed, delayed care from long appointment times, a conceptual measure of delayed access to care, results in poor health outcome ^40^, which further leads to an increased cost of care and poverty ^14,36^. Therefore, it can be conceptualized that individuals with COPD fall into a virtual loop characterized by delayed access to care, increased cost of care, worsening poverty, and worsening health outcomes, ultimately culminating in mortality ^14^.

Our study has some limitations. The data were drawn from an interview survey, and respondents self-reported their health status. The likelihood of recall bias, therefore, cannot be eliminated. Misclassification of asthma as COPD may have occurred, although this possibility is undermined as the NHIS has separate questions for asthma, emphysema, and chronic bronchitis. Separating these conditions limited the risk of misclassification, but it does not eliminate it. This study, being an ecological study, makes causal inferences impossible. Although we controlled for smoking – a major risk factor for COPD, we did not assess the influence of other genetic factors, occupational exposures, and other related chronic diseases due to a large amount of missing data or absent variables that define these factors. However, we conceptualize that these factors will further increase the odds of functional limitation and not weaken the observed effect. Despite these limitations, this study is strengthened by its robust pool of eleven-year survey-weighted data, making inferential bias less likely. It is one of the few studies that highlight the impact of healthcare access in the care of individuals with COPD and the role of poverty in worsening functional limitations. This study serves as a useful policy instrument in the continued advocacy for improving the health indices of individuals with COPD.

## Conclusion

Among individuals with COPD, functional limitations are associated with a lack of healthcare coverage, delayed appointment, unaffordable care, and lack of a usual place for care. The odds of functional limitation is further heightened among those who are poor. This study presents areas of further research, as other measures of health outcome such as the relationship of measures of health care access and mortality pattern among individuals with COPD is yet to be explored in the literature.

## Data Availability

IPUMS-NHIS data 2008 to 2018 restricted to patients with COPD and functional limitation

